# New plasma LC-MS/MS assays for the quantitation of beta-amyloid peptides and identification of apolipoprotein E proteoforms for Alzheimer’s disease risk assessment

**DOI:** 10.1101/2023.11.20.23298532

**Authors:** Darren M. Weber, Jueun C. Kim, Scott M. Goldman, Nigel J. Clarke, Michael K. Racke

## Abstract

**Objectives:** Early detection of Alzheimer’s disease (AD) represents an unmet clinical need. Beta-amyloid (Aβ) plays an important role in AD pathology, and the Aβ42/40 peptide ratio is a good indicator for amyloid deposition. In addition, variants of the *APOE* gene are associated with variable AD risk. Here we describe the development and validation of high-throughput liquid chromatography-tandem mass spectrometry (LC-MS/MS) assays for plasma Aβ40 and Aβ42 quantitation, as well as apolipoprotein E (ApoE) phenotype determination as a surrogate for *APOE* genotype.

**Methods:** Aβ40 and Aβ42 were simultaneously immunoprecipitated (IP) from plasma, proteolytically digested, and quantitated by LC-MS/MS. ApoE proteoform status was qualitatively assessed by targeting tryptic peptides from the ApoE2, ApoE3, and ApoE4 proteoforms. Both assays were validated according to CLIA guidelines.

**Results:** Within-run precision was 1.8 to 4.2% (Aβ40), 1.9 to 7.2% (Aβ42), and 2.6 to 8.3% (Aβ42/40 ratio). Between-run precision was 3.5 to 5.9% (Aβ40), 3.8 to 8.0% (Aβ42), and 3.3 to 8.7% (Aβ42/40 ratio). Both Aβ40 and Aβ42 were linear from 10 to 2,500 pg/mL. Identified ApoE proteoforms had 100% concordance with *APOE* genotypes.

**Conclusion:** We have developed a precise, accurate, and sensitive high-throughput LC-MS/MS assay for plasma Aβ40, Aβ42, and proteoforms of ApoE.

## Introduction

Early detection of Alzheimer’s disease (AD) has become critically important with the United States Food and Drug Administration’s recent approval of the first disease-modifying treatments for AD using monoclonal antibodies targeting removal of various aggregates of Aβ from the brain [1]. Positron emission tomography (PET) and measurement of cerebrospinal fluid (CSF) beta-amyloid 42 (Aβ42) [2, 3] are methods that have been used as entry criteria for clinical trials and/or as outcome measures for -disease-modifying AD treatments, but are costly and invasive [4].

Blood-based biomarkers have shown promise as a low-cost, noninvasive means of detecting amyloid pathology, and several studies in plasma have demonstrated that the ratio of Aβ42 to Aβ40 beta-amyloid peptides (Aβ42/40) is inversely correlated with amyloid burden [5-9]. In addition, polymorphisms in the apolipoprotein E gene (*APOE*) have been shown to be a major genetic risk factor for late-onset AD, with individuals with the *APOE4* allele having a 3- to 15-fold increase in their odds of developing AD compared with more common *APOE3* allele [10]. In addition, recent studies have shown that the combination of the plasma Aβ42/40 with age and *APOE4* status, determined by identifying apolipoprotein E proteoforms (ApoE), can identify amyloid pathology with higher accuracy than Aβ42/40 alone [11].

Mass spectrometry (MS)-based methods have demonstrated higher accuracy in identifying amyloid deposition compared to immunoassays [5], most likely due to better analytical specificity and less susceptibility to matrix effects. However, MS-based platforms are considered complex and costly, and not as accessible as automated immunoassays. Most employ sample preparation workflows involving peptide enrichment using immunoprecipitation (IP) coupled with detection of peptides using costly high-resolution MS instruments [12, 13]. An antibody-free, low resolution MS method was recently reported, but relies on ion mobility for background suppression and microfluidics, making it difficult to assess robustness and throughput [14].

Given the superiority of MS-based methods for AD biomarker detection and quantitation, and the need for better accessibility in AD testing, Quest Diagnostics has developed 2 liquid chromatography-tandem mass spectrometry assays (LC-MS/MS assays). Full automation and multiplexing enable assay robustness and high throughout, and costs are minimized by using a high-sensitivity but low-resolution triple quadrupole MS. The first assay simultaneously quantitates plasma levels of Aβ40 and Aβ42 (Quest AD-Detect™, Beta-Amyloid 42/40 Ratio). The second assay identifies ApoE proteoforms/phenotype as a surrogate for *APOE* geneotype (Quest AD-Detect™, Apolipoprotein E Isoform). Herein, we describe their development and validation.

## Materials and methods

### Specimen collection

Blood specimens were collected by venipuncture into 10 mL tubes containing EDTA as anticoagulant, kept on ice (<1 hour) until centrifugation at 1200 rcf for 12 minutes at room temperature. Aliquots of plasma (0.5 mL) were transferred into polypropylene tubes and stored at -80⁰C until analysis.

### Preparation of plasma Aβ calibrators, internal standards, and quality control samples

Full-length Aβ40 and Aβ42, were purchased from rPeptide (Watkinsville, GA USA). Quantitative amino acid analysis (AAA) was used to confirm purity and check stated peptide content of the Aβ40 and Aβ42 peptides, which was adjusted accordingly as previously described [15].

Aβ40 and Aβ42 calibrators were prepared by dissolving each peptide in 6 M urea containing 10% (v/v) stripped plasma (Golden West, Temecula, CA) in 10 mM PBS to a final concentration of 1 mg/mL. Both peptides were combined and subsequently diluted to a final concentration of 50 ng/mL in 6 M urea containing 10% stripped plasma. Calibrators were frozen at -80⁰C in 200 µL aliquots in 2.0 mL Protein LoBind^®^ Eppendorf tubes (Eppendorf, Hamburg, Germany) that had pre-treated to prevent non-specific analyte loss as previously described [16].

Quality control (QC) samples were prepared as described above using a separate lot Aβ40 and Aβ42 peptides. A total of 4 QC samples were used for the assay. The low QC consisted of a neat plasma pool, and the high QC consisted of the same plasma pool spiked with Aβ40 and Aβ42. Two additional QC samples were prepared by spiking Aβ40 and Aβ42 into stripped plasma to mimic a probable AD specimen (Aβ42/40 ∼ 0.120; AD-QC) and a probable non-AD specimen (Aβ42/40 ∼0.300; nonAD-QC). All QC samples were aliquoted into Protein LoBind Eppendorf tubes and stored at -80⁰C until needed.

Isotopically-labeled Aβ40 (uniformly-labeled 13C, 15N) and Aβ42 (uniformly-labeled 15N) internal standard (IS) were purchased from rPeptide (Watkinsville, GA USA). The IS mix was prepared as described above to a final concentration of 10 ng/mL, and 200 µL were aliquoted in 2.0 mL Protein LoBind® Eppendorf tubes and stored at -80⁰C until needed.

### Plasma Aβ40 and Aβ42 sample preparation

On the day of the assay, frozen calibrators, QC samples, IS, and patient plasma specimens were thawed. Once thawed, samples were processed using a fully automated method on Hamilton Star MicroLab liquid handler (Hamilton, Reno, NV, USA), configured to process two 96 deep-well plates per batch. First, 100 µL of 1% Tween-20/CHAPS (v/v) was added to each 96 deep-well polypropylene plate, followed by addition of 500 µL of calibrators, QC samples, and patient plasma specimens, and 500 µL of 10 mM PBS. Next, IS was added to all samples to a final concentration of 500 pg/mL for both Aβ40 and Aβ42. Samples were then mixed for 20 minutes at room temperature at 1,100 rpm. Finally, Aβ40 and Aβ42 were enriched from plasma by immunoprecipitation (IP) using 5 µg of biotinylated monoclonal anti-Aβ17-24 antibody (clone 4G8, Biolegend, San Diego, CA) conjugated to Dynabeads™ MyOne™ Streptavidin T1 magnetic beads (Invitrogen, Waltham, MA). After a 2-hour IP binding step, the beads were washed twice with 1 mL of 0.01% Tween-20 in 10 mM PBS followed by 2 washes with 1 mL of 0.12 M ammonium bicarbonate (AMBIC). Next, 250 µL of 6 M urea was added to each sample and mixed for 10 minutes at 1,400 rpm at room temperature. To each sample was added 250 µL of 0.12 M AMBIC, followed by 2 µg of endoproteinase Lys-C (Santa Cruz Biotechnology, Dallas, TX), which yielded Aβ29-40 (amino acid sequence GAIIGLMVGGVV) and Aβ29-42 (amino acid sequence GAIIGLMVGGVVIA). Protein digestion proceeded at 40⁰C with mixing at 1,200 rpm. After a 70-minute incubation, the digestion reaction was quenched by adding 100 µL of 10% ammonium hydroxide, and samples were concentrated and desalted using a micro-elution mixed-mode anion exchange solid phase extraction (SPE) plate (Waters, Milford, MA). Samples were loaded onto the SPE plate, washed with 10% acetonitrile, and eluted with 120 µL of 60% acetonitrile with 5% formic acid. Samples were further diluted with 120 µL of water prior to LC-MS/MS analysis.

### Quantification of plasma Aβ40 and Aβ42 by LC-MS/MS

The 96-well sample plate containing digested Aβ40 and Aβ42 was placed into an LC-autosampler set to a temperature of 4⁰C, and 70 µL was injected onto a XBridge Protein BEH 300 Å C4 column (Waters, Milford, MA) heated to 55⁰C. Analytical separation was achieved using a Transcend Vanquish TLX-4 TurboFlow UPLC (Thermo Fisher Scientific, Waltham, MA, USA) in a staggered 4-column configuration to facilitate high throughput. Both peptides were resolved using a 16-minute gradient at a flow rate of 0.6 mL/min of solvent A (water with 0.15% formic acid) and solvent B (acetonitrile with 0.15% formic acid) with a 2-minute acquisition window. Detection was achieved using a Thermo Scientific TSQ Altis Plus Triple Quadrupole MS (Thermo Fisher Scientific, Waltham, MA, USA) operated in multiple reaction monitoring (MRM) mode. Optimized collision energies and RF voltage values were determined by direct infusion of the digested peptides. Three unique transitions for both Aβ40 and Aβ42, as well as their respective IS, were monitored. Parent/production ion masses, collision energies, and RF lens voltages are summarized in **Table 1**. Ion source and MRM scan parameters are summarized in **Supplemental material, Table 2**.

**Table 1:** Precursor/product ion masses, collision energies, and RF lens values for Aβ40, Aβ42, and their respective internal standards. [Place near the section titled “Quantification of plasma Aβ40 and Aβ42 by LC-MS/MS” in the Methods section]

| Analyte and Peptide Sequence | Precursor Ion (m/z) | Product Ion (m/z) | Collision Energy (V) | RF Lens (V) |
| --- | --- | --- | --- | --- |
| Aβ40: GAIIGLMVGGVV | 1085.6 <sup>(+1)</sup> | 812.4 (b9) | 32 | 155 |
|  |  | 869.4 (b10) | 33 | 155 |
|  |  | 968.5 (b11) | 28 | 155 |
| Aβ40 IS: GAIIGLMVGGVV<sup>a</sup> | 1146.7 <sup>(+1)</sup> | 858.6 (b9) | 32 | 155 |
|  |  | 918.6 (b10) | 33 | 155 |
|  |  | 1023.7 (b11) | 28 | 155 |
| Aβ42: GAIIGLMVGGVVIA | 1269.8 <sup>(+1)</sup> | 968.5 (b11) | 35 | 194 |
|  |  | 1067.6 (b12) | 35 | 194 |
|  |  | 1180.6 (b13) | 32 | 194 |
| Aβ42 IS: GAIIGLMVGGVVIA<sup>b</sup> | 1294.8 <sup>(+1)</sup> | 987.6 (b11) | 35 | 194 |
|  |  | 1092.7 (b12) | 35 | 194 |
|  |  | 1205.8 (b13) | 32 | 194 |

<sup>a</sup> uniformly-labeled 13C/15N; <sup>b</sup> uniformly-labeled 15N

**Table 2:**
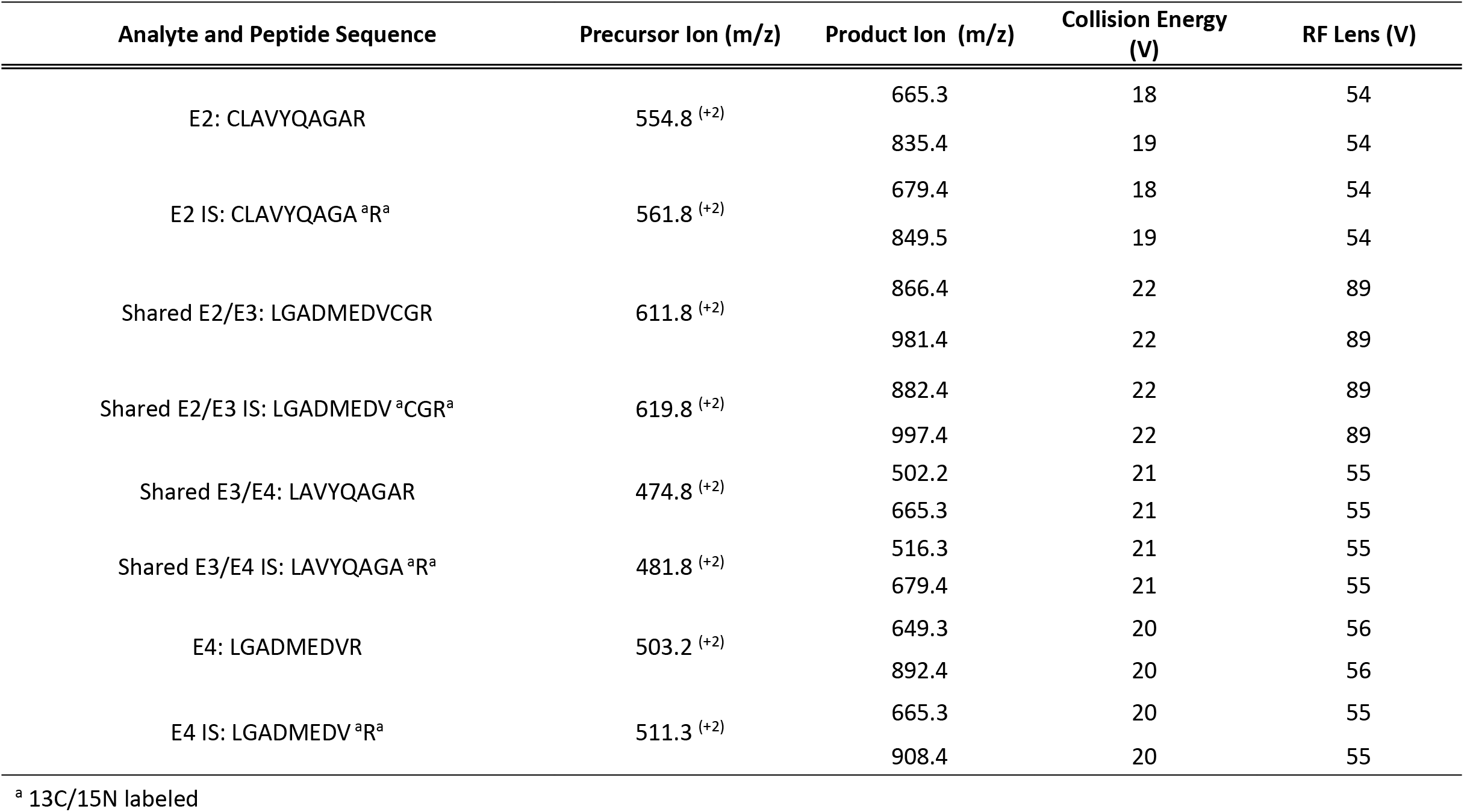
Precursor/product ion masses, collision energies, and RF lens values for ApoE peptides and their respective internal standards. [Place near the section titled “Qualitative determination of ApoE proteoforms by LC-MS/MS” in the Methods section]

Each 96-well plate consisted of an 8-point calibration curve spanning the linear range (10 to 2,500 pg/mL) for both Aβ40 and Aβ42, a set of front QCs, a maximum of 79 plasma specimens, and a set of back QCs dispersed throughout the plate so to bracket patient specimens. The ratio of the peak area of the analyte to the internal standard was used to calculate the concentrations from the standard curve using TraceFinder Clinical Research v5.1 software (Thermo Fisher Scientific, Waltham, MA, USA). A weighted quadratic model (1/x) was used for generation of the standard curves while ignoring the origin. The ratio of Aβ42 to Aβ40 (Aβ42/40) was determined by taking the back-calculated value for Aβ42 and dividing it by the back-calculated value for Aβ40. The total allowable error (TAE) for Aβ40 and Aβ42 was 30%. An example chromatogram from a patient plasma specimen is shown in **Supplemental material, Figure S1**.

### Qualitative determination of ApoE proteoforms by LC-MS/MS

On the day of the assay, frozen QC samples, IS, and patient plasma specimens were thawed. Once thawed, samples were processed using a fully automated method on Hamilton Star MicroLab liquid handler configured to process two 96 deep-well plates per batch. Samples (25 µL) were diluted in 125 µL of 0.12 M AMBIC containing 15 mM dithiothreitol, 0.5% sodium deoxycholate (DOC), and 1 µg of winged internal standard (Biosynth, Gardner, GA, USA) and incubated for 20 min at 45⁰C. Next, iodoacetamide was added to a final concentration of 20 mM. After a 10 min incubation at 45⁰C, 3 µg of trypsin was added and the samples were proteolytically digested for 2 hours at 55⁰C. Post digestion, samples were acidified with formic acid, centrifuged for 10 minutes at 2,250 rcf to pellet the DOC, and desalted and concentrated using an Agilent BondElut^®^ C18 SPE plate (Agilent, Santa Clara, CA, USA). ApoE plasma digests were loaded onto the SPE plate, washed with 0.1% formic acid, and eluted with 500 µL of 80% acetonitrile with 0.1% formic acid. Samples were evaporated under heated nitrogen and resuspended in 200 µL of 5 % acetonitrile with 0.1% formic acid prior to LC-MS/MS analysis.

Unique tryptic peptides to the ApoE2 (CLAVYQAGAR) and ApoE4 (LGADMEDVR) proteoforms, as well as shared peptides between the ApoE2 and ApoE3 proteoforms (LGADMEDVCGR), and the ApoE3 and ApoE4 proteoforms (LAVYQAGAR), were targeted for identification. Analytical separation was achieved using a Transcend Vanquish TLX-4 TurboFlow UPLC (Thermo Fisher Scientific, Waltham, MA, USA) using a 4-column setup. Peptides were chromatographically resolved using an Agilent Poroshell 120 EC-C18 column. All peptides were resolved using a 6-minute gradient and a flow rate of 0.6 mL/min of solvent A (water with 0.15% formic acid) and solvent B (acetonitrile with 0.15% formic acid) with a 1-minute acquisition window. Detection was achieved using a Thermo TSQ Altis Triple Quadrupole MS operated in MRM mode (**Supplemental material, Table S2**). Optimized collision energies and RF voltage values were determined by direct infusion of the digested peptides. Two unique transitions for each peptide and IS were monitored. Parent/production ion masses, collision energies, and RF lens voltages are summarized in **Table 2**. ApoE phenotypes were determined by the presence or absence of the unique and the shared peptides, as summarized in **Table 3**. An example chromatogram from a patient plasma specimen with an ApoE2/ApoE4 phenotype, showing the presence of all 4 monitored ApoE proteoform-specific peptides, is shown in **Supplemental material, Figure S2**. Peak area intensity thresholds and the relationship between analyte and IS retention times were used to identify ApoE phenotype peptides using TraceFinder Clinical Research v5.1 software (Thermo Fisher Scientific, Waltham, MA, USA).

**Table 3:** ApoE peptide signatures for proteoform determination. [Place near the section titled “Qualitative determination of ApoE proteoforms by LC-MS/MS” in the Methods section]

| **ApoE Phenotype** | **ApoE2 Peptide:**<br>**CLAVYQAGAR** | **Shared ApoE2/ApoE3 Peptide:**<br>**LGADMEDVCGR** | **Shared ApoE3/ApoE4 Peptide:**<br>**LAVYQAGAR** | **ApoE4 Peptide:**<br>**LGADMEDVR** |
| --- | --- | --- | --- | --- |
| E2/E2 | Present | Present | - | - |
| E2/E3 | Present | Present | Present | - |
| E2/E4 | Present | Present | Present | Present |
| E3/E3 | - | Present | Present | - |
| E3/E4 | - | Present | Present | Present |
| E4/E4 | - | - | Present | Present |

### Validation

Assay precision (within-run and between-run), analytical measurement range (AMR), analytical sensitivity (limit-of-blank [LOB], limit-of-detection [LOD], and limit-of-quantification [LOQ]), interference testing, carryover, and stability were conducted according to CLSI guidelines [17-19]. A detailed description of each validation study is described in the Supplemental material.

## Results

### Plasma Aβ40 and Aβ42

### Precision

Within-run and between-run assay precision results for Aβ40, Aβ42, and the Aβ42/40 ratio are presented in **Table 4**. Within-run imprecision ranged from 1.8 to 4.2% for Aβ40, 1.9 to 7.2% for Aβ42, and 2.6 to 8.3% for the Aβ42/40 ratio. Total imprecision (between-run) ranged from 3.5 to 5.9% for Aβ40 (average imprecision of 4.3%), 3.8 to 8.0% for Aβ42 (average imprecision of 5.6%), and 3.3 to 8.7% for the Aβ42/40 ratio (average imprecision of 5.5%).

**Table 4:** Precision results for plasma Aβ40, Aβ42, and the Aβ42/40 ratio. [Place near the section titled “Precision” under “Plasma Aβ40 and Aβ42” in the Results section]

| Aβ40 | N | Mean (pg/mL) | Within Run (Intra-assay) | | Between Run (Inter-assay) | |
| --- | --- | --- | --- | --- | --- | --- |
|  |  |  | SD | CV | SD | CV |
| S1 | 25 | 163.3 | 4.0 | 2.4% | 6.9 | 4.2% |
| S2 | 25 | 520.1 | 12.8 | 2.5% | 18.2 | 3.5% |
| S3 | 25 | 619.2 | 20.6 | 3.3% | 23.1 | 3.7% |
| S4 | 25 | 1091.9 | 20.0 | 1.8% | 46.7 | 4.3% |
| S5 | 25 | 177.3 | 7.4 | 4.2% | 10.5 | 5.9% |

| Aβ42 | N | Mean (pg/mL) | Within Run (Intra-assay) | | Between Run (Inter-assay) | |
| --- | --- | --- | --- | --- | --- | --- |
|  |  |  | SD | CV | SD | CV |
| S1 | 25 | 40.0 | 2.9 | 7.2% | 3.2 | 8.0% |
| S2 | 25 | 64.4 | 2.5 | 3.8% | 2.5 | 3.8% |
| S3 | 25 | 188.1 | 7.3 | 3.9% | 9.7 | 5.1% |
| S4 | 25 | 902.5 | 17.3 | 1.9% | 39.9 | 4.3% |
| S5 | 25 | 36.9 | 2.1 | 5.7% | 2.6 | 7.0% |

| Aβ42/40 | N | Mean | Within Run (Intra-assay) | | Between Run (Inter-assay) | |
| --- | --- | --- | --- | --- | --- | --- |
|  |  |  | SD | CV | SD | CV |
| S1 | 25 | 0.245 | 0.020 | 8.3% | 0.021 | 8.7% |
| S2 | 25 | 0.124 | 0.005 | 4.0% | 0.006 | 4.7% |
| S3 | 25 | 0.304 | 0.014 | 4.6% | 0.018 | 5.8% |
| S4 | 25 | 0.827 | 0.022 | 2.6% | 0.027 | 3.3% |
| S5 | 25 | 0.207 | 0.009 | 4.5% | 0.011 | 5.2% |

### Analytical sensitivity

For Aβ40, the LOB was 1.9 pg/mL and the LOD was 3.5 pg/mL. For Aβ42, the LOB was 1.6 pg/mL and the LOD was was 2.8 pg/mL The LOQ for both Aβ40 and Aβ42 was 10 pg/mL.

### Analytical measurement range

Aβ40 was linear from 25 to 1,000 pg/mL and Aβ42 was linear from 10 to 500 pg/mL, with a coefficient of determination of ≥0.99 for both peptides (**Figure 1**). For Aβ40, the CVs across the 6 dilution levels ranged from 1.2 to 4.1%, and the difference between the expected and observed values ranged from -7.2 to 4.7%. For Aβ42, the CVs across the 6 dilution levels ranged from 1.2 to 7.1%, and the difference between the expected and observed values ranged from -2.2 to 6.2%.

**Figure 1:**
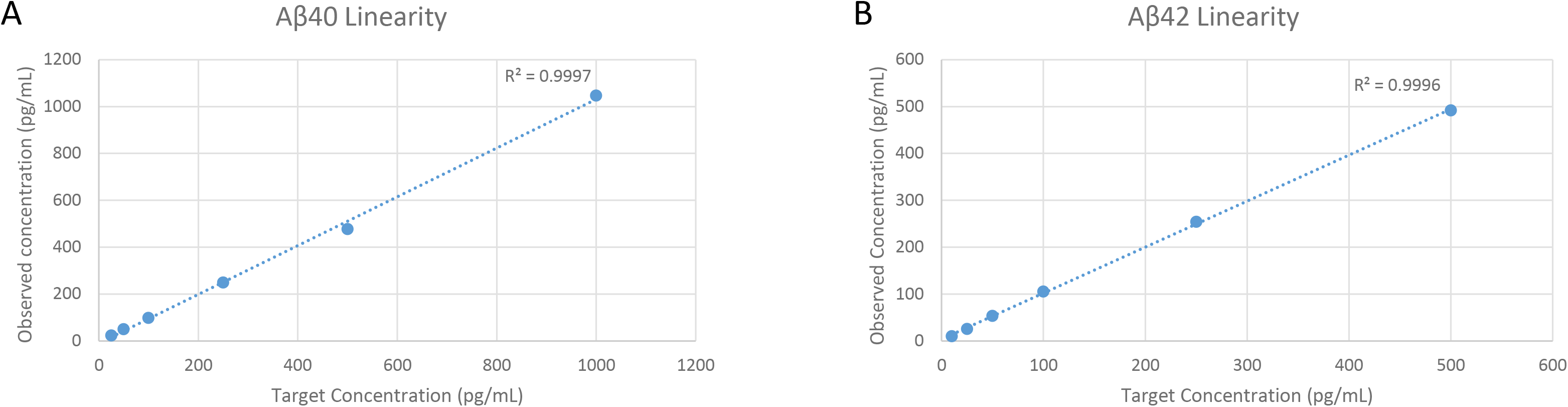
Linearity for (A) Aβ40 and (B) Aβ42. [Place near the section titled “Analytical Measurement Range” under “Plasma Aβ40 and Aβ42” in the Results section]

### Accuracy and recovery

Recoveries for spiked patient plasma specimens ranged from 93.2 to 98.9% for Aβ40, and 94.0 to 97.5% for Aβ42. Recoveries for low and high patient plasma mixes were from 98.1 to 101.5% for Aβ40, and from 97.2 to 103.0% for Aβ42.

### Interference

No interference was observed for lipemia and icterus at the tested concentrations for both Aβ40 and Aβ42 (**Supplemental material, Table 1**). Low (40 mg/dL) and moderate (80 mg/dL) amounts of hemolysis were acceptable for both Aβ40 and Aβ42. However, high levels of hemolysis (≥ 800 mg/dL) showed a 29% decrease in the recovery of Aβ40 and a 31% decrease in the recovery of Aβ42. Despite this, the effects of gross hemolysis were mitigated by the Aβ42/40 ratio, which showed an average recovery of 96.4% compared to baseline values. The method showed no evidence of ion suppression for Aβ40 and Aβ42.

### Specimen stability

Stability at room temperature was 8 hours for Aβ40, 4 hours for Aβ42, and 4 hours for the Aβ42/40 ratio (**Figure 2A**). After 24 hours of storage at room temperature, the average recovery for Aβ42 declined by nearly half (56%), whereas Aβ40 showed an average recovery of 78%. Aβ40, Aβ42, and the Aβ42/40 ratio were stable up to 5 days refrigerated (**Figure 2B**), at least 32 days when stored frozen (**Figure 2C**), and at least 5 months at ultralow temperature (**Figure 2D**). Aβ40, Aβ42, and the Aβ42/40 ratio were stable for up to 5 freeze/thaw cycles, with average recoveries ranging from 93.6 to 101.9% for Aβ40, 86.2 to 101.4% for Aβ40, and 89.2 to 105.0% for the Aβ42/40 ratio.

**Figure 2:**
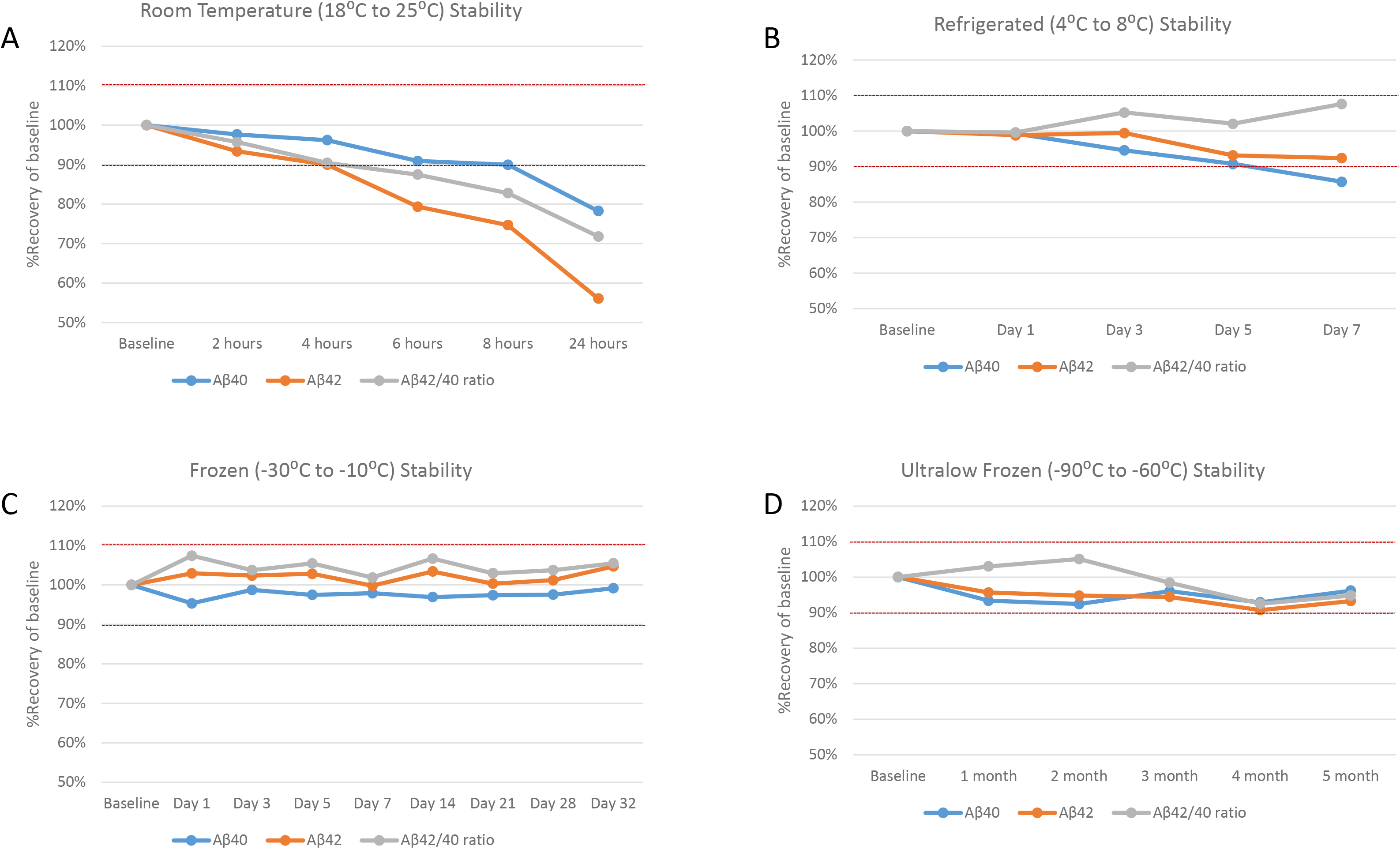
Average percent recovery for Aβ40, Aβ42, and the Aβ42/40 ratio under various storage conditions: (A) room temperature (18⁰C to 25⁰C); (B) refrigerated (4⁰C to 10⁰C); (C) frozen (-30⁰C to -10⁰C); and (D) ultra-low frozen (-90⁰C to -60⁰C). [Place near the section titled “Specimen Stability” under “Plasma Aβ40 and Aβ42” in the Results section]

Post-digested and extracted samples showed stability up to 2 days when stored at 4⁰C in the instrument’s autosampler.

### Carryover

The assay showed no evidence of carryover for Aβ40 and Aβ42.

### Plasma ApoE proteoform

### Repeatability and accuracy

Repeatability was 100% for all 6 tested ApoE phenotype samples analyzed over 5 separate days, with 5 replicates of each sample tested. Accuracy studies showed 100% concordance with the PCR genotype results (**Table 5**).

**Table 5:** Accuracy of ApoE proteoforms with APOE genotypes. [Place near the section titled “Repeatability and accuracy” under “Plasma ApoE Proteoforms” in the Results section]

| **Genotype** | **Expected # of Samples** | **Actual # of Samples** | **PPA (%)** |
| --- | --- | --- | --- |
| *APOE 2/2* | 1 | 1 | **100%** |
| *APOE 2/3* | 20 | 20 | **100%** |
| *APOE 2/4* | 3 | 3 | **100%** |
| *APOE 3/3* | 141 | 141 | **100%** |
| *APOE 3/4* | 72 | 72 | **100%** |
| *APOE 4/4* | 13 | 13 | **100%** |

PPA, positive percent agreement

### Interference, stability, and carryover

The accurate determination of the 6 ApoE phenotypes was not affected by the presence of hemolysis, lipemia, or icterus at the concentrations tested. All 6 ApoE phenotypes were shown to be stable for at least 14 days at room temperature and refrigerated, at least 3 months frozen (-30⁰C to -10⁰C), and for least 3 months frozen at ultralow temperature (-90⁰C to -70⁰C). All 6 phenotypes were also shown to be stable for at least 5 freeze/thaw cycles. Digest and extracted samples were shown to be stable for at least 5 days when stored at 4⁰C. The assay showed no evidence of carryover for the 6 tested ApoE phenotypes.

## Discussion

We report the validation of a fully automated, multiplexed method for the simultaneous quantification of Aβ40 and Aβ42 by IP-LC-MS/MS and the qualitative determination of ApoE proteoforms by LC-MS/MS. Both assays showed excellent analytical characteristics. Linearity studies and precision studies for Aβ40, Aβ42, and the Aβ42/40 ratio showed good reproducibility, with average overall analytical variability being <6% and detection limits well below peptide levels observed in plasma. The ApoE proteoform assay demonstrated 100% concordance with PCR genotyping. In addition, both assays utilize a high-throughput multiplexed HPLC system coupled to a triple quadrupole MS, a low-resolution instrument found in most clinical diagnostic laboratories. This contrasts with similar approaches that rely on specialized high-resolution MS instrumentation and/or single-plex HPLC systems [12, 14]. With this method, 158 plasma Aβ40 and Aβ42 samples and 176 plasma ApoE samples can be processed per batch.

Blood-based biomarkers for AD have several advantages over CSF, most notably specimen accessibility. However, several challenges exist in developing sensitive, specific, and robust assays for blood biomarkers for AD. While CSF is in continuous contact with the brain, resulting in higher levels of certain protein biomarkers, the fraction of these biomarkers that enter the bloodstream is much lower [20]. In fact, we found that concentrations of Aβ40 and Aβ42 were roughly 100-fold lower in plasma than in CSF [16]. To overcome these challenges, we employed a protein IP step in order to enrich for both Aβ peptides and decrease matrix complexity. This fully automated protein IP step exhibited high reproducibility and allowed for detection of Aβ40 and Aβ42 concentrations well below expected plasma values. Employing protein IP also eliminated any adverse effects due to endogenous interferents.

Additionally, blood-based biomarkers present several preanalytical factors that must be considered. The time periods between blood collection and centrifugation and storage, as well as post-centrifugation plasma storage conditions, are important [21, 22] and were evaluated as part of our assay validation study. Although using the ratio of Aβ42 to Aβ40 can mitigate some preanalytical effects compared with using Aβ42 alone [23], rates of loss of the Aβ40 and Aβ42 peptides can differ. We observed a noticeable reduction in the levels of Aβ42 after 4 hours at room temperature relative to Aβ40, similar to findings in other studies [24]. Average recoveries for Aβ42 decreased roughly twice as fast as Aβ40 (-20% vs. <-10%, **Figure 2**) when stored at room temperature, suggesting that Aβ42 is more susceptible to proteolytic cleavage, aggregation, or absorptive loss under these conditions. However, when stored at 4⁰C, stability for both Aβ peptides increased to 5 and 7 days for Aβ42 and Aβ40, respectively. Unlike at room temperature, the average recoveries of Aβ40 and Aβ42 changed proportionally at 4⁰C, resulting in the Aβ42/40 ratio being stable for at least 7 days.

An additional challenge for the clinical application Aβ42/Aβ40 ratio is the lack of certified reference materials for either analyte. We, and others, have used quantitative AAA to adjust for variable peptide content in supplier-provided powders [12]. Although vendors may produce peptides with similar levels of purity, variable peptide content can result in lot-to-lot discrepancies in calibrator ratios. In contrast to many laboratories, we routinely perform these measurements independent of the supplier [15].

Apolipoprotein E is the most abundant apolipoprotein in the central nervous system, and individuals with the *APOE4* allele have elevated Aβ deposition in the brain compared to individuals with the most common *APOE3* allele [10] and are at a higher risk of developing amyloid-related imaging abnormalities (ARIA) when treated with anti-amyloid monoclonal antibody therapeutics [25, 26]. Given the clinical significance of ApoE in AD pathology and therapy, methods that can accurately identify ApoE proteoforms are needed. In keeping with previous studies [12], our qualitative LC-MS/MS assay demonstrated 100% a concordance with PCR genotyping. Importantly, the low-volume specimen requirements of each assay (500 µL for Aβ and 25 µL for ApoE) permit the use of both AD-Detect assays on the same patient specimens. This provides an opportunity to combine assessment of Aβ status and AD and ARIA risks using a single plasma specimen from each patient.

## Conclusion

We have described the development and validation of LC-MS/MS assays that accurately quantify plasma-derived Aβ40, Aβ42, and Aβ42/40 ratio, and accurately identify ApoE proteoforms. In contrast to previous studies, the assays do not rely on specialized high-resolution MS instrumentation, thus reducing assays costs, and/or single-plex HPLC systems, thus enhancing assay throughput and robustness. Clinical studies that evaluate the diagnostic performance of each assay have been completed, including assessment of the association of the number of *APOE4* alleles with the Aβ42/40 ratio, and will be presented elsewhere.

## Data Availability

All data produced in the present study are available upon reasonable request to the authors.

## Abbreviations

Aβ: beta-amyloid
APOE: apolipoprotein E
TAE: total allowable error
MRM: multiple reaction monitoring
DOC: sodium deoxycholate
IS: internal standard

## Acknowledgements

The authors thank Thomas Lynn and Nicolas Valdivia for programming and assistance with the automation. We also thank the University of Florida for providing genotyping specimens and results and Jiyeon Ahn and Chris McKernan for their assistance with the analytical validation, and Dr. Steven Taylor for his review and comments of the manuscript.

