## Supplemental material for "New plasma LC-MS/MS assays for the quantitation of beta-amyloid peptides and identification of apolipoprotein E proteoforms for Alzheimer’s disease risk assessment"

IRB APPROVED
AS MODIFIED
Sep 17, 2022

### RESEARCH SUBJECT INFORMATION AND CONSENT FORM

**TITLE:** Banking of Adult Human Specimens at Quest Diagnostics Nichols Institute

**PROTOCOL NO.:** BR13-002
WCG IRB Protocol #20121940

**SPONSOR:** Quest Diagnostics Nichols Institute

**INVESTIGATOR:** Michael McPhaul, MD
33608 Ortega Highway
San Juan Capistrano, California 92675
United States

**STUDY-RELATED PHONE NUMBER(S):** Michael McPhaul, MD
949-728-44702
949-702-5805 (24 hours)


Maritza Ayllon
Daniel Quezada
Karla Arroyo
Vanessa Centeno
949-728-4051
949-728-4757 (fax)


This consent form may contain words that you do not understand. Please ask the study doctor or the study staff to explain any words or information that you do not clearly understand. You may take home an unsigned copy of this consent form to think about or discuss with family or friends before making your decision.

#### Purpose of the Study

The purpose of this study is to build a specimen bank of samples. Banked samples will be used for reference intervals, specimen stability studies, specimen comparisons, and other validation needs.

The tests done on your specimens are for research purposes only and will not affect any medical treatment that you may be undertaking. Any samples collected will not have any markings or labels to identify the sample with your name. However, samples may be marked with the date and time that the sample was collected.

IRB APPROVED
AS MODIFIED
Sep 17, 2022

**Procedures**
You will be asked to complete a health history case report form which includes a list of any prescription and over-the-counter drugs, herbs, and vitamins that you may be taking. You will also be asked about foods you have recently eaten, as well as any significant health issues.

If you agree to be in this research a small amount of your blood will be drawn from a vein in your arm. You also may be asked to provide specimens of your urine, stool, and other body fluids.

You may be asked to provide specimens up to twice weekly, provided you remain within the collection requirements. If you give your consent to participate in this research, it will be good for one year. You will be asked to sign a new consent form each year if you decide to continue providing specimens to Quest Diagnostics. You may also be contacted after specimen donation for follow-up questions such as delayed side effects (i.e., bruising, pain or residual bleeding) or changes to their medical history when needed.

**Genetic Testing**
Your specimens may be used for genetic testing. These studies may include, but are not limited to: biochemical studies, molecular genetic markers, and molecular genetic sequencing. If your samples will be used in one of these studies, you will be asked to sign a separate genetic consent document.

**Risks**
Drawing blood may cause pain, bruising, lightheadedness, or, on rare occasions, infection. Also, as with any research study, there is the possibility of loss of confidentiality of your information.

**New Findings**
You will be told about any new information that might change your decision to participate in this study. You may be asked to sign a revised consent form if this occurs.

**Benefits**
Although you will receive no direct benefits from being in this study, participation in this research study may lead to improved medical testing for the community.

**Costs**
There are no costs associated with participating in this study.

IRB APPROVED
AS MODIFIED
Sep 17, 2022

**Payment for Participation**

Participant research subjects are compensated for their time and inconvenience in participating in this study. Subjects will be reimbursed as follows:

| | | |
|---|---|---|
| Blood | | |
| Finger stick Sample (one stick) | | $20.00 |
| Venipuncture (arm) ≤20 ml | | $20.00 |
| Venipuncture (arm) >20 ml | | $1.00 / ml |
| Capillary Collection (one device) | | $30.00 |
| Swab/Saliva (one sample) | | $20.00 |
| Urea Breath Collection (1 device) | | $30.00 |
| Urine | | |
| Single Sample | | $20.00 |
| 24-hour Collection | | $75.00 |
| Stool | | |
| One Collection | | $55.00 |
| Other Non-invasive Collections (one sample) | | $20.00 |
| Missed stick | (up to 2 missed sticks) | $20.00 each |
| Multiple samples (e.g. OGTT) | | $20.00 each |
| Tasso and YourBio Self Collection | | $125.00 for one device evaluation and $250.00 for both |

If you are an employee of Quest Diagnostics, you will be reimbursed through your paycheck approximately 30 days after your participation in the study. If you are not a Quest employee, you will receive a check approximately 2 months after your participation. Please ask the study staff if you have questions about when you will be paid.

**Compensation for Injury**

In the event of physical injury directly resulting from the laboratory procedures used to obtain the sample according to the study plan, Quest Diagnostics Nichols Institute will provide compensation for reasonable medical expenses. There are no plans to provide any other type of compensation.

**Alternatives**

Your alternative is to not participate in this study.

**Source of Funding**

Funding for this research study is provided by Quest Diagnostics, Nichols Institute.

IRB APPROVED
AS MODIFIED
Sep 17, 2022

**Privacy**

Quest Diagnostics strictly adheres to issues of the right to privacy (including applicable federal and state laws regarding privacy). All electronic study documents will be stored on password protected computers with limited access, and all paper study documents will be store in locked cabinets. Furthermore, Quest Diagnostics will not disclose information resulting from this testing to any third party including family members, employers, potential employers, or health insurance companies.

In the event that a critical result is discovered during the testing of your sample, our medical director would like to send you a copy of the outlying result so that you may consult with a physician at your discretion. However, if you prefer to not receive this information, it will not be provided.

A Federal law called the Genetic Information Nondiscrimination Act (GINA) provides some protection for your genetic information. This law generally will protect you in the following ways:

- Health insurance companies and group health plans may not request your genetic information collected in this research.
- Health insurance companies and group health plans may not use your genetic information when making decisions regarding your eligibility or premiums.
- Employers with 15 or more employees may not use your genetic information collected in this research when making a decision about your employment.

However, this Federal law does not protect you against genetic discrimination by companies that sell life insurance, disability insurance, or long-term care insurance.

**Scientific Communication of Results**

Quest Diagnostics may report these test results in scientific formats, such as scientific / medical meetings, scientific correspondence or peer reviewed papers submitted to specialized journals. However, your identity will never be disclosed. Any identity disclosure, in the form of written information or picture(s) must be previously authorized by you in a special signed document.

**Storage of the Specimen**

Banked samples will be stored until they are used up or destroyed.

**Voluntary Participation/Withdrawal**

Quest Diagnostics management does not urge, influence, or encourage anyone who works for the company to take part in a research study. Your participation in this study is completely voluntary. You may withdraw from the study at any time and for any reason. Your decision to not participate in the study, or a decision on your part to withdraw from the study, will have no effect whatsoever on your (or any family member's) employment status at Quest Diagnostics. You may refuse to participate, or you may withdraw from the study at any time without penalty, prejudice, or loss of benefits to which you are entitled.

If you withdraw from the study, any remaining specimens will be discarded, and no additional information will be gathered.

Your participation in this study may be stopped at any time by the study doctor or the sponsor without your consent for any of the following reasons:

- if it is in your best interest;
- or for any other reason.

**Questions**

If you have any questions about this study, the Principal Investigator, Michael McPhaul, MD is available to provide that information.

949-728-4702 (office hours)
949-702-5805 (24 hours)

Any study coordinator can also be contacted with study questions.
949-728-4051(office hours)


- if you have any questions concerning your participation in this study,
- if at any time you feel you have experienced a research-related injury, or
- if you have questions, concerns or complaints about the research.

If you have questions about your rights as a research subject **or if you have questions, concerns or complaints about the research, you may contact:**

WCG IRB
1019 39th Avenue SE Suite 120
Puyallup, Washington 98374-2115
.

WCG IRB is a group of people who perform independent review of research.

WCG IRB will not be able to answer some study-specific questions, such as questions about appointment times.  However, you may contact WCG IRB if the research staff cannot be reached or if you wish to talk to someone other than the research staff.

Do not sign this consent form unless you have had a chance to ask questions and have received satisfactory answers to all of your questions.

You will be given a copy of this signed and dated consent form and the Experimental Subject's Bill of Rights to keep for your records.

IRB APPROVED
AS MODIFIED
Sep 17, 2022

**Consent**

I certify that I have been informed and consent that my specimen can be used for assay validations and research at Quest Diagnostics.

It has been explained that I may withdraw my participation in this study at any time with no adverse consequences. This study will be performed at no charge.

All my questions about this research study and my participation in it have been answered to my satisfaction. I voluntarily agree to be in this study.

By signing this consent form, I have not given up any of my legal rights.

Printed Subject Name Date

________________________________________ _______________________

Signed Subject Name

________________________________________

If a critical result is discovered during testing of my sample, I would (please initial)

_____ like to receive a copy of the result

_____ prefer not to be sent the result

We would like permission to contact you to see if you would like to donate more samples. (please initial)

______Yes, please contact me if you would like me to donate samples again.

______No, please do not contact me to request additional samples.

Signature of Person Conducting Informed Consent Discussion Date

________________________________________ _______________________

IRB APPROVED
AS MODIFIED
Sep 17, 2022

### AUTHORIZATION TO USE AND DISCLOSE INFORMATION FOR RESEARCH PURPOSES

Federal regulations give you certain rights related to your health information. These include the right to know who will be able to get the information and why they may be able to get it. The study doctor must get your authorization (permission) to use or give out any health information that might identify you.

**What information may be used and given to others?**
If you choose to be in this study, the study doctor will get personal information about you. This may include information that might identify you. The study doctor may also get information about your health including:

- Research records
- Records about your study visits
- Information obtained during this research about laboratory tests

**Who may use and give out information about you?**
Information about your health may be used and given to others by the study doctor and staff. They might see the research information during and after the study.

The study doctor may provide the date and time of sample collection with your study samples to researchers.

**Who might get this information?**
Your information may be given to the sponsor of this research. “Sponsor” includes any persons or companies that are working for or with the sponsor, or are owned by the sponsor.

Information about you and your health which might identify you may be given to:

- The U.S. Food and Drug Administration (FDA)
- Department of Health and Human Services (DHHS) agencies
- WCG IRB

**Why will this information be used and/or given to others?**
Information about you and your health that might identify you may be given to others to carry out the research study. The sponsor will analyze and evaluate the results of the study. In addition, people from the sponsor and its consultants will be visiting the research site. They will follow how the study is done, and they will be reviewing your information for this purpose.

The information may be given to the FDA. The information may be used to meet the reporting requirements of governmental agencies.

The results of this research may be published in scientific journals or presented at medical meetings, but your identity will not be disclosed.

The information may be reviewed by WCG IRB. WCG IRB is a group of people who perform independent review of research as required by regulations.

**What if I decide not to give permission to use and give out my health information?**
By signing this consent form, you are giving permission to use and give out the health information listed above for the purposes described above. If you refuse to give permission, you will not be able to be in this research.

**May I review or copy the information obtained from me or created about me?**
You have the right to review and copy your health information. However, if you decide to be in this study and sign this permission form, you will not be allowed to look at or copy your information until after the research is completed.

**May I withdraw or revoke (cancel) my permission?**
This permission will be good until December 31, 2050.

You may withdraw or take away your permission to use and disclose your health information at any time. You do this by sending written notice to the study doctor. If you withdraw your permission, you will not be able to continue being in this study.

When you withdraw your permission, no new health information which might identify you will be gathered after that date. Information that has already been gathered may still be used and given to others. This would be done if it were necessary for the research to be reliable.

**Is my health information protected after it has been given to others?**
If you give permission to give your identifiable health information to a person or business, the information may no longer be protected. There is a risk that your information will be released to others without your permission.

IRB APPROVED
AS MODIFIED
Sep 17, 2022

**Authorization:**
I have been given the information about the use and disclosure of my health information for this research study. My questions have been answered.

I authorize the use and disclosure of my health information to the parties listed in the authorization section of this consent for the purposes described above.

Printed Subject Name Date

____________________________________ ______________________

Signed Subject Name

____________________________________

Signature of Person Conducting
Informed Consent Discussion Date

____________________________________ ______________________
