## Supplemental material for "New plasma LC-MS/MS assays for the quantitation of beta-amyloid peptides and identification of apolipoprotein E proteoforms for Alzheimer’s disease risk assessment"

### Plasma Aβ40, Aβ42, Aβ42/40 ratio analytical validation

#### Precision

Assay precision was conducted using a nested design to evaluate both within-run (repeatability) and between-run precision (1). A total of 5 precision study samples were tested over 25 days, with 1 run per day and 2 replicates per tested sample (25x1x2), using a one lot of reagents and one instrument. Mean, standard deviation (SD) and coefficient of variance (CV) were calculated for Aβ40, Aβ42, and the Aβ42/40 ratio to determine within-run and between-run precision. Study samples included: S1) Low QC; S2) AD-QC; S3) nonAD-QC; S4) High QC; S5) patient plasma pool. Acceptability for within-run and between-run precision was TAE/3, or 10%.

#### Analytical Sensitivity

The limit of blank (LOB) study was conducted to determine the highest measurement result likely to be observed from a blank sample (2). The LOB study used 2 lots of sample diluent analyzed on one instrument over one day, with 25 replicates of each blank sample. The LOB was evaluated for both Aβ40 and Aβ42 using the following formula, which is a modification of the formula suggested in CLSI EP17A2:

$$\text{LOB} = \text{mean of blank} + 2(\text{SD})$$

The limit of detection (LOD) study consisted of the same sample diluent specimens used in the LOB study. The LOD was evaluated for both Aβ40 and Aβ42 using the following formula, which is a modification of the formula suggested in CLSI EP17A2:

$$\text{LOD} = \text{mean of blank} + 4(\text{SD})$$

The limit of quantification (LOQ) study was conducted to determine the lowest concentration that can accurately be quantified. The LOQ study was evaluated using a 3x5x5 design, with 3 LOQ specimens (10, 25, and 50 pg/mL for both Aβ40 and Aβ42) analyzed over 5 days, with 5 replicates per day. The mean, SD, and CV were calculated for each LOQ sample for both Aβ peptides, and the LOQ was determined as the lowest concentration that at which CV was ≤20%.

**Analytical Measurement Range**

Analytical measurement range (AMR) studies were conducted to evaluate the linearity of the test (3). A high concentration sample was prepared by spiking Aβ40 and Aβ42 into a pooled plasma specimen. The high concentration sample was then serial diluted to create 5 additional levels, and each level was analyzed in triplicate. Mean, SD, CV, and % recovery of expected values were evaluated for each level. Back-calculated values for each level were plotted, and linearity was assessed by regression analysis. Linearity was acceptable if the deviation of each level did not exceed TAE/4, or 7.5% of the expected values, and coefficient of determination ($R^2$) was ≥0.99.

Additionally, linearity was evaluated by recovery on dilution of plasma specimens. Five separate plasma specimens were analyzed neat and diluted 1 to 2 with sample diluent. Each tested sample was then analyzed in triplicate within the same run, and observed Aβ40 and Aβ42 values were compared to the expected values. Recovery on dilution was deemed acceptable if the observed values were within 20% of the expected values.

**Accuracy and Recovery**

Accuracy was evaluated by spiking known concentrations of recombinant Aβ40 and Aβ42 into 5 separate plasma specimens. Each plasma specimen was analyzed neat and spiked with 50, 250, and 600 pg/mL of Aβ40 and Aβ42. All samples were then analyzed in triplicate within the same run. The percent recovery was determined by comparing the observed concentrations of the spiked samples to the expected concentrations. Recovery within 20% of expected values was deemed acceptable.

Recovery of mixed samples was evaluated by analyzing a plasma pool with high levels of Aβ40 and Aβ42 and a plasma pool with low levels of Aβ40 and Aβ42. Both the high pool and low pool samples were then mixed in a 1 to 1 ratio to make a "high-low" pool. Next, the "high-low" pool was mixed with equal volumes of the high pool ("high-mix") and low pool ("low-mix") to create 2 additional plasma sample pools. All samples were then analyzed in triplicate within the same run. The percent recovery was determined by comparing the observed concentrations of the spiked samples to the expected concentrations. Recovery within 20% of expected values was deemed acceptable.

**Interference**

Potential interference from endogenous compounds was evaluated in three different patient plasma samples. Hemolysis, icterus, and lipemia was tested at 3 different levels summarized in **Table 3**. Each tested plasma sample was analyzed neat and spiked with a low, moderate, or high concentration of each interferent. All samples were analyzed in triplicate. Interference was considered insignificant if the deviation between neat and spiked sample was ≤TEA/4, or 7.5%.

Ion suppression was evaluated by post-column infusion of the proteolytic C-terminal peptides of both Aβ40 and Aβ42. Chromatograms were assessed visually for a decrease in the total ion chromatogram (TIC) at the retention time of each analyte.

**Specimen stability**

Sample stability was assessed by storing 8 plasma specimens at room temperature (18 ºC to 25ºC; baseline, 2, 4, 6, 8, and 24 hours), 5 plasma specimens refrigerated (2ºC to 8ºC; baseline, 1, 3, 5, and 7 days), 5 plasma specimens frozen (-30ºC to -10ºC, baseline, 1, 3, 5, 7, 14, 21, 28, and 32 days), and 5 plasma specimens frozen at ultralow (baseline, 1, 2, 3, 4, and 5 months). Except for the ultralow stability samples, after each time point, samples were frozen at -80 ºC. All samples were subsequently thawed and analyzed on the same day. Freeze–thaw stability was assessed for 5 freeze–thaw cycles on 5 different patient serum pools. Extracted sample stability was assessed at 0- and 2-days post-extraction. Baseline values were compared to the observed values at each stability time point to determine a percent recovery. Stability was assessed for Aβ40, Aβ42, and the Aβ42/40 ratio. Stability was acceptable if the deviation between baseline values and stability time points/storage conditions was ≤ TAE/3, or 10%.

**Carryover**

Carryover was assessed by analyzing 4 matrix blank samples followed by 8 replicates of a pooled plasma specimen overspiked with 1,000 pg/mL of Aβ40 and Aβ42. Next, 8 matrix blank samples were analyzed and evaluated for evidence of carryover.

**Plasma ApoE Proteoform Validation**

All validation studies were performed using the 6 common ApoE phenotypes: ApoE2/E2, ApoE2/E3, ApoE2/E4, ApoE3/E3, ApoE3/E4, and ApoE4/E4. All phenotype samples, except for the ApoE2/E2 and ApoE4/E4, were prepared using pooled human plasma. The ApoE2/E2 phenotype sample and the ApoE4/E4 phenotype sample was prepared by spiking recombinant ApoE2 protein or ApoE4 protein into sample diluent, respectively.

**Plasma ApoE Proteoform Repeatability**

Repeatability (precision) studies were performed by analyzing 5 replicates of each phenotype sample over 5 separate days using one instrument and one operator. Criteria for acceptance was 100% concordance with the expected phenotype result.

**Plasma ApoE Proteoform Accuracy**

Accuracy for ApoE was evaluated by concordance with genotyping. A comparison study of 250 plasma specimens, previously genotyped at the University of Florida using real-time PCR restriction length polymorphism, was analyzed over 4 runs, with one run per day. Results from our ApoE proteoform assay were compared to the PCR results. Acceptable criteria were established as 100% agreement between the 2 assays.

**Plasma ApoE Proteoform Interference**

Potential interference from endogenous compounds was evaluated in all 6 ApoE phenotype specimens. Hemolysis, icterus, and lipemia was tested at 3 different levels summarized in **Table 3**. Each tested plasma sample was analyzed neat and spiked with a low, moderate, or high concentration of each interferent. All samples were analyzed in triplicate. Acceptability criteria was established as 100% agreement with the expected phenotype result.

Ion suppression was evaluated by post-column infusion of targeted ApoE tryptic peptides.

**Plasma ApoE Proteoform Stability**

Sample stability was assessed by storing each ApoE phenotype specimen at room temperature (18 ºC to 25ºC; baseline, 1, 3, 5, and 7 days), refrigerated (2ºC to 8ºC; baseline, 1,

3, 5, 7, and 14 days), frozen (-30ºC to -10ºC, baseline, 1, 3, 5, 7, 14, 21, 26, and 31 days), and ultra-low frozen (-90ºC to -70ºC, baseline, 7, 14, 21, 30 days, and 3 months). After each time point, samples were frozen at -80 ºC. All samples were subsequently thawed and analyzed on the same day. Freeze–thaw stability was assessed for 5 freeze-thaw cycles on each ApoE phenotype specimen. Extracted sample stability was assessed at 0-, 1-, 2-, and 3-days post-extraction. Acceptance criteria for storage condition and stability time was determined as 100% concordance with the expected ApoE phenotype.

**Plasma ApoE Proteoform Carryover**

Carryover was assessed by analyzing 4 matrix blank samples followed by 4 replicates of each ApoE phenotype specimen spiked with recombinant ApoE2 or ApoE4 to be roughly 3 times the average ApoE concentration. Next, 8 matrix blank sample were analyzed and evaluated for evidence of carryover.

**Table S1**: List of potential endogenous interferents and their tested concentrations

| Interferent | Tested Low Concentration | Tested Moderate Concentration | Tested High Concentration |
|---|---|---|---|
| Hemolysis (washed and lysed RBC) | ~40 mg/dL | ~80 mg/dL | ~800 mg/dL |
| Icterus (unconjugated bilirubin) | 0.5 mg/dL | 1 mg/dL | 10 mg/dL |
| Lipemia (Intralipid) | 0.5 mg/dL | 1 mg/dL | 10 mg/dL |

Abbreviations: RBC = red blood cells

**Table S2:** HESI source settings and MRM scan parameters for Aβ40 and Aβ40 and ApoE.

| **HESI Source Parameters** | **Aβ40 and Aβ42** | **Apolipoprotein E** |
|---|---|---|
| Spray Voltage (V) | 4000 | 3500 |
| Sheath Gas | 50 | 50 |
| Aux Gas | 5 | 5 |
| Sweep Gas | 1 | 1 |
| Ion Transfer Tube Temp ($^{o}$C) | 380 | 350 |
| Vaporizer Temp ($^{o}$C) | 450 | 400 |
| **MRM Scan Parameters** | **Aβ40 and Aβ42** | **Apolipoprotein E** |
| Polarity | Positive | Positive |
| Q1 Resolution (FWHM) | 1.2 | 0.7 |
| Q3 Resolution (FWHM) | 1.2 | 1.2 |
| CID Gas (mTorr) | 2 | 2 |

Abbreviations: HESI = heated electrospray ionization; MRM = multiple reaction monitoring; FWHM= full-width half mass

**Figure S1**: LC-MS/MS chromatograms for Aβ40 and Aβ40 IS (top pane) and Aβ42 and Aβ42 IS (bottom pane) from a patient plasma specimen.

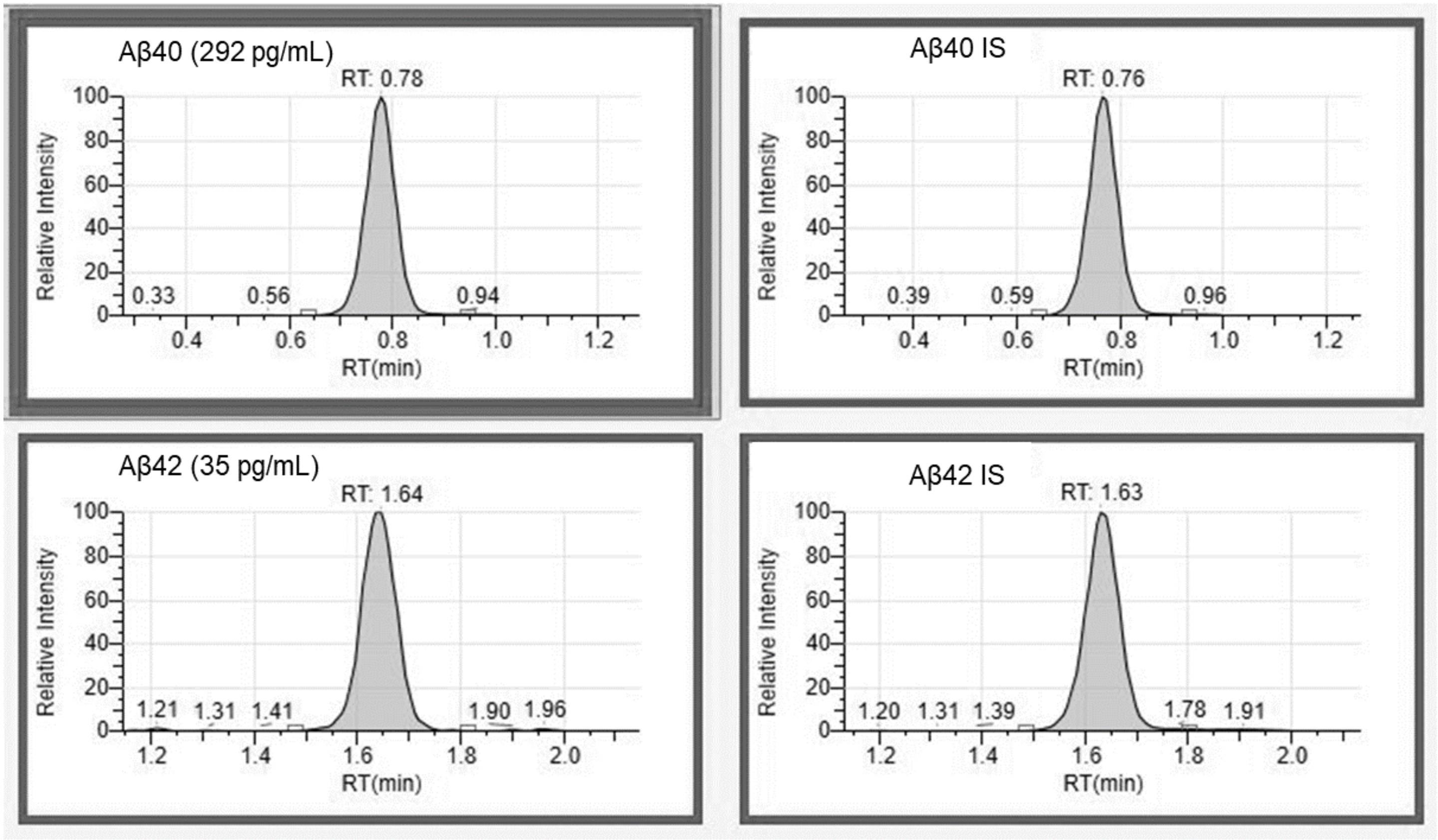

**Figure S2**: LC-MS/MS chromatogram from a patient plasma specimen with an ApoE2/ApoE4 phenotype, showing the presence of all 4 targeted ApoE proteoform peptides.

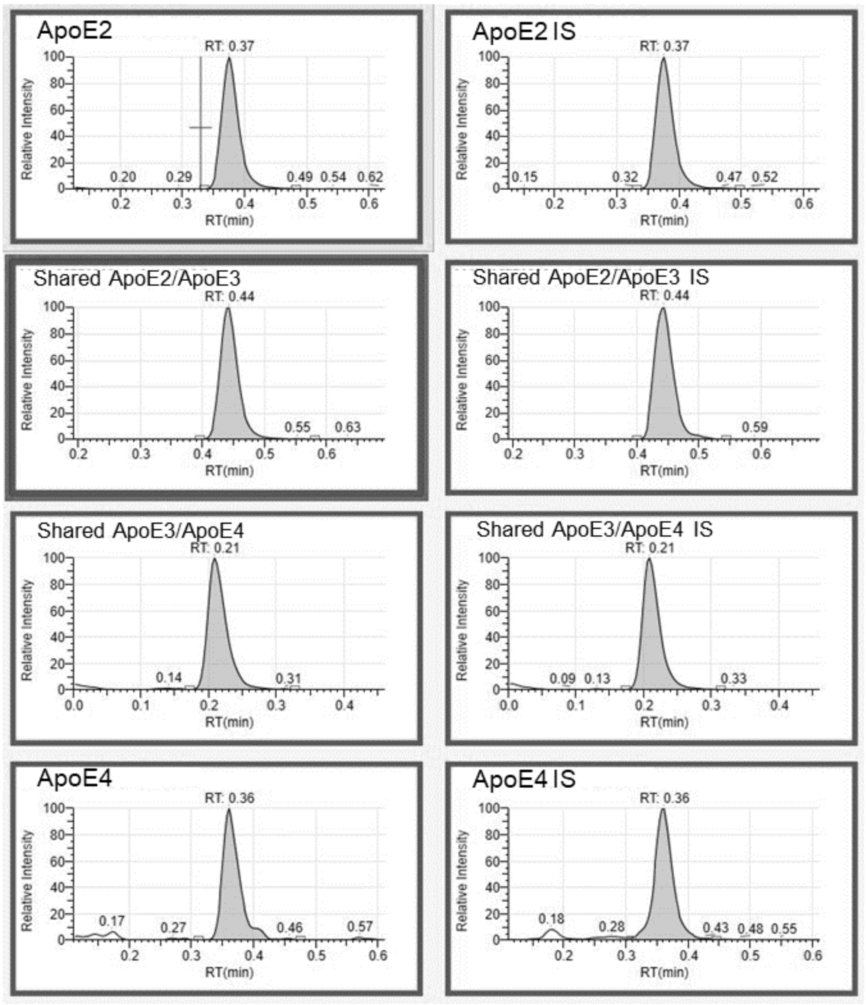


### References


1. CLSI. Evaluation of Precision of Quantitative Measurement Procedures; Approved Guideline—Third Edition. CLSI document EP05-A3. Wayne, PA: Clinical and Laboratory Standards Institute. 2014.
2. CLSI. Evaluation of Detection Capability Implementation Guide. 1st ed. CLSI implementation guide EP17-Ed2-IG. Clinical and Laboratory Standards Institute. 2021.
3. CLSI. User Verification of Linearity Implementation Guide. 1st ed. CLSI implementation guide EP06-Ed2-IG. Clinical and Laboratory Standards Institute. 2022.
